# ECG-Guided Pre-Screening of Family Members for Hypertrophic Cardiomyopathy

**DOI:** 10.64898/2026.06.19.26354464

**Authors:** Ali Lootah, Sandar Min, Meena Fatah, Robert M Hamilton, Seema Mital

**Affiliations:** Department of Pediatrics, The Hospital for Sick Children, University of Toronto, Toronto, Ontario Canada; Genetics and Genome Biology Program, The Hospital for Sick Children, Toronto, Ontario, Canada; Ted Rogers Centre for Heart Research, University of Toronto, Toronto, Ontario, Canadas

**Keywords:** Hypertrophic cardiomyopathy, ECG, echocardiography, family screening

## Abstract

**Background:** Current clinical guidelines recommend serial ECG and echocardiographic surveillance for first-degree relatives of probands with Hypertrophic Cardiomyopathy (HCM).

**Objectives:** To evaluate the accuracy and validity of ECG alone as a pre-screening tool for the diagnosis of HCM and to develop a random forest (RF) model for HCM phenotype prediction.

*Method:* Pediatric relatives of primary HCM probands attending the cardiomyopathy screening program at The Hospital for Sick Children were included from 1993 till 2025. Subjects were followed until last follow-up censored at phenotype conversion. ECGs were classified as normal or abnormal based on predefined parameters. Associations between binary ECG variables and HCM phenotype were assessed using Phi (φ) coefficient. A Random Forest classifier was developed using significant ECG variables (70:30 training:test split) and evaluated using precision, recall, specificity, negative predictive value, F1 score and AUROC. feature importance was assessed using SHAP analysis. Variables with impact of >5% were included in a simplified model which was evaluated by repeating performance metrics and externally validated in a healthy cohort.

**Results:** 350 screened relatives (44% female, mean follow-up 6.8 ± 4.8 years) were included. At baseline 13% (46\350) were phenotype-positive for HCM. 9 subjects converted during the surveillance. Thirteen ECG variables were significantly associated with phenotype-positive HCM and were included in the full random forest model. Four variables had >5% impact (Left ventricular hypertrophy, right ventricular hypertrophy, T-wave inversion and ST-segment depression) and were included in a simplified model which maintained high specificity (93% vs 97%), negative predictive value (97% vs 93%) and AUROC (90% vs, 96%). The simplified model classified 83% subjects as phenotype-negative with eight being false-negative, all of whom developed an abnormal ECG in a mean of 1 year and none had an interim adverse cardiac event. The simplified model was evaluated in an independent healthy cohort of 153 school age subjects and correctly identified 98% as phenotype-negative with 100% NPV.

**Conclusion:** ECG abnormalities were strongly associated with phenotype-positive status. A simplified ECG-based random forest model using four ECG variables demonstrated high specificity and negative predictive value for identifying phenotype-negative subjects. If prospectively validated, this could reduce the need for concurrent echocardiographic screening by up to 83% per encounter, lowering screening burden and cost.

## INTRODUCTION

Hypertrophic cardiomyopathy (HCM) is a leading cause of cardiomyopathy and of sudden cardiac death in young athletes^1^. ECG abnormalities are common in HCM, but their independent predictive value in family screening is uncertain^2^. Current guidelines recommend combined rather than tiered ECG and echocardiographic screening for surveillance of first-degree relatives^3^. We developed a machine learning model incorporating ECG features and evaluated its performance as a pre-screening test to identify individuals at low risk of phenotype-positive HCM in whom echocardiographic screening could potentially be deferred.

## METHODS

This retrospective single-center study included pediatric relatives (<18 years old) of primary HCM probands evaluated through the cardiomyopathy screening program at the Hospital for Sick Children (Toronto, Canada) between 1993 and 2025. Exclusion criteria were missing ECGs or echocardiograms, non-sinus rhythm, secondary cardiomyopathy, and lack of follow-up. The study was approved by the institutional Research Ethics Board and waiver of informed consent was obtained.

Serial ECGs and echocardiograms from initial screening to last follow-up censored at HCM phenotype conversion were analyzed. Phenotype-positivity was defined echocardiographically as an interventricular septal or left ventricular posterior wall thickness z-score of ≥2.5 at a single encounter, or two consecutive values between 2.0–2.5 at sequential encounters^3^. ECGs were manually reviewed by a single operator blinded to echocardiographic phenotype (A.L). ECGs were defined as abnormal if morphological, conduction, depolarization, or repolarization parameters^2^ fell outside normal pediatric reference ranges as defined in published literature^4–6^.

Associations between ECG variables and phenotype-positive HCM were assessed. Categorical variables were summarized as frequencies (%), and continuous variables as means ± standard deviation. Group comparisons were performed using Chi-square or Fisher exact test for categorical variables, and Student’s t-test for continuous variables. Collinearity among ECG features and HCM phenotype was assessed using pairwise Phi (φ) correlation coefficients.

A random forest classifier model using ECG parameters at last follow-up to classify HCM phenotype was trained and tested using a 70:30 train-test split. Model performance was evaluated using area under the receiver operating characteristic curve (AUROC), precision or positive predictive value (PPV), sensitivity, specificity, F1-score, and negative predictive value (NPV). SHapley Additive exPlanations (SHAP) analysis quantified each variable’s contribution to model prediction. A simplified model incorporating the top-performing variables was subsequently constructed, and model performance was evaluated in the HCM testing dataset. It was further validated in an independent healthy control dataset that included previously published ECG data from clinically healthy children with no underlying heart disease^7^.

## RESULTS

Three hundred fifty pediatric relatives from 136 HCM families were included (44% female; baseline age 6.4±4.6 years). At baseline, 13% were phenotype-positive, increasing to 15.6% over 6.8±4.8 years of follow-up. 71% phenotype-positive, and 19% phenotype-negative individuals had abnormal ECGs at baseline (**Figure 1a**). Nine subjects (2.6%) showed phenotype conversion during follow-up. Thirteen ECG variables were associated with phenotype status (p<0.05) (**Figure 1a**). All variables were retained for model development as phi (φ) coefficient demonstrated no significant collinearity (all <0.7).

**Figure 1.**
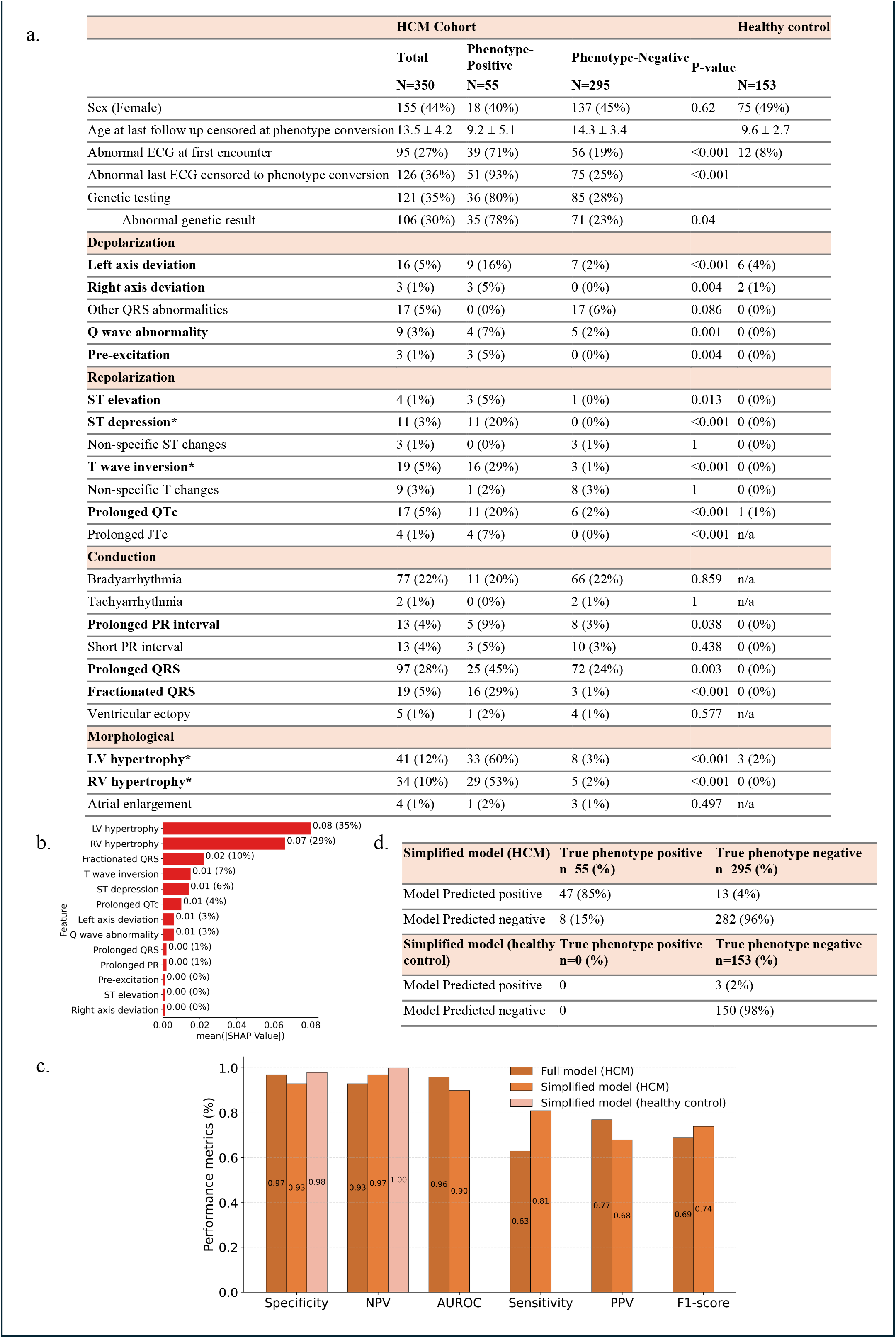
**a)** Clinical characteristics of hypertrophic cardiomyopathy (HCM) screening cohort (stratified by phenotype status), and healthy control cohort. P-values represent comparison between HCM phenotype subgroups. Bolded variables were included in the full model; asterisk indicates variables included in the simplified model. **b)** Mean absolute SHapley Additive exPlanations (SHAP) values demonstrating the relative contribution of each ECG variable to HCM phenotype prediction. Higher SHAP values indicate greater contribution to model prediction with percentage contribution shown in parentheses. **c)** Clustered column chart showing the performance metrics of the full (13-variable) and simplified (4-variable) model. Random Forest models in the testing dataset at last follow-up (dark brown and orange respectively) and in the healthy control dataset (pink). **d)** Classification results of the simplified ECG model in the overall HCM cohort at last follow-up and healthy control cohort. Values represent the absolute number and percentage of individuals identified as phenotype positive or negative compared with true phenotype status. *LV, left ventricular; RV, right ventricular; SHAP, SHapley Additive exPlanations; NPV, negative predictive value; AUROC, area under the receiver operating characteristic curve; PPV, positive predictive value; QTc, Corrected QT interval; JTc, Corrected JT interval*

Subjects were randomly divided into training (70%, n=245) and testing (30%, n=105) datasets. A random forest classifier model predicting HCM phenotype using the thirteen statistically significant ECG variables was developed using the training dataset (full model). SHAP analysis identified 5 variables with ≥5% contribution to model prediction, (LV hypertrophy, RV hypertrophy, T-wave inversion, ST-segment depression and QRS fractionation). QRS fractionation was excluded due to inconsistent reporting by cardiologists, and the remaining 4 variables were used to develop a simplified model (**Figure 1b**).

Both models demonstrated an AUROC, specificity, and NPV ≥90% for identifying phenotype-negative individuals when applied to the testing set (**Figure 1c**). Given its reliance on more readily identifiable ECG features, the simplified model was further evaluated in the overall HCM cohort to assess misclassification rates. The model correctly identified 96% (282/295) of phenotype-negative and 85% (47/55) of phenotype-positive individuals at the last encounter censored at phenotype conversion (**Figure 1d**). If prospectively validated, the model could reduce the numbers of individuals undergoing echocardiographic screening from 350 to 60 individuals (83% reduction). However, 8 phenotype-positive individuals (15%) were misclassified as phenotype-negative by the ECG model. All subsequently developed an abnormal ECG after a median follow-up of 1 year and none experienced a serious interim adverse event.

The simplified model was also evaluated in an independent healthy cohort (n=153; 49% female; age 9.6±2.7 years) of whom 8% had ECG abnormalities. Model performance remained high, correctly identifying 98% (150/153) of individuals as phenotype-negative, with only 3 individuals misclassified as phenotype-positive (**Figure 1d**).

## DISCUSSION

In pediatric relatives of HCM probands, ECG abnormalities were common among phenotype-positive but infrequent in phenotype-negative individuals. Using machine-learning, four ECG variables (LV hypertrophy, RV hypertrophy, T-wave inversion and ST-segment depression), emerged as the strongest predictors of HCM phenotype. A model incorporating these variables demonstrated high accuracy for identifying phenotype-negative individuals both in HCM family members and in an independent healthy cohort.

Current practice guidelines recommend that all family members undergo routine echocardiographic screening every 1-3 years even though^3^, as our data show, only 13% were phenotype-positive at first evaluation and another 2.6% became positive over more than 6 year follow-up. If validated prospectively as a first-line screening test, the simplified ECG model could potentially reduce the need for concurrent echocardiographic screening by 83%. The model misclassified 8 patients as phenotype-negative without adverse consequences of delayed diagnosis.

These findings suggest that a tiered approach to HCM screening using ECG as a pre-screening test may be considered in order to reduce the burden and costs of concurrent echocardiographic screening in individuals with a low likelihood of being phenotype-positive. While use of AI approaches in ECG screening is gaining traction, we relied on conventional ECG features that are routinely measured clinically and therefore more readily applicable in clinical decision making and for use as a decision support tool. Prospective validation in an independent HCM cohort is needed to determine the potential utility in reducing costs and burden of echocardiographic surveillance in pediatric HCM family members.

## Data Availability

All data produced in the present study are available upon reasonable request to the authors

